# Disposable N95 Masks Pass Qualitative Fit-Test But Have Decreased Filtration Efficiency after Cobalt-60 Gamma Irradiation

**DOI:** 10.1101/2020.03.28.20043471

**Authors:** Avilash Cramer, Enze Tian, Sherry H. Yu, Mitchell Galanek, Edward Lamere, Ju Li, Rajiv Gupta, Michael P. Short

## Abstract

The current COVID-19 pandemic has led to a dramatic shortage of masks and other personal protective equipment (PPE) in hospitals around the globe [1]. One component of PPE that is in particular demand are disposable N95 face masks. To alleviate this, many methods of N95 mask sterilization have been studied and proposed with the hope of being able to safely reuse masks [2]. Two major considerations must be made when re-sterilizing masks: (1) the sterilization method effectively kills pathogens, penetrating into the fibers of the mask, and (2) the method does not degrade the operational integrity of the N95 filters.

We studied Cobalt-60 (^60^Co) gamma irradiation as a method of effective sterilization without inducing mask degradation. Significant literature exists supporting the use of gamma radiation as a sterilization method, with viral inactivation of SARS-CoV reported at doses of at most 10 kGy [3], with other studies supporting 5 kGy for many types of viruses [4]. However, concerns have been raised about the radiation damaging the fiber material within the mask, specifically by causing cross-linking of polymers, leading to cracking and degradation during fitting and/or deployment [5, 6].

A set of 3M 8210 and 9105 masks were irradiated using MIT’s ^60^Co irradiator. Three masks of each type received 0 kiloGray (kGy), 10 kGy and 50 kGy of approximately 1.3 MeV gamma radiation from the circular cobalt sources, at a dose rate of 2.2kGy per hour.

Following this sterilization procedure, the irradiated masks passed a OSHA Gerson Qualitative Fit Test QLFT 50 (saccharin apparatus) [7] when donned correctly, performed at the Brigham and Women’s Hospital, in a blinded study repeated in triplicate. However, the masks’ filtration of 0.3 *µ*m particles was significantly degraded, even at 10 kGy.

These results suggest against gamma, and possibly all ionizing radiation, as a method of disposable N95 sterilization. Even more importantly, they argue against using the qualitative fit test alone to assess mask integrity.

## Introduction

An N95 mask, when fitted correctly, blocks 95% of 0.3 micron particles, and provides an important barrier against disease transmission as part of a healthcare provider’s personal protective equipment (PPE). Unfortunately, a shortage of N95 masks globally amidst the COVID-19 pandemic [1] has forced healthcare providers in hard hit areas to use a single mask for hours or days at time- or simply go without adequate personal protective equipment (PPE) [8, 9].

Given the ease of access to N95 masks during normal times, much remains unknown about the efficacy of various sterilization mechanisms on disposable masks since there is little impetus to reuse these masks outside of major disaster and pandemic events. Several methods of sterilizing N95 masks between uses are currently being studied across the world. The advantages of these methods and concerns regarding their use are summarized in Table 1. Vaporized hydrogen peroxide and EtO treatments in particular are described in detail by Viscusi et al (2009) [2]; UV treatment procedures are described in Darnell & Taylor (2006) and Kumar et al (2015) [10].

**Table 1.** Advantages and concerns of proposed N95 sterilization techniques.

| Technique | Advantages | Concerns |
| --- | --- | --- |
| Gamma radiation ($^{60}\text{Co}$, $^{137}\text{Cs}$, linear accelerators) | Gamma radiation shown to work well for viral inactivation; used in food sterilization<br>High range of gamma radiation enables batch sterilization for higher throughput, large volumes<br>High range of gamma radiation also enables sterilization inside biosafety container, easing transport and handling | High doses of gamma radiation can degrade some polymers; irradiation process may take time depending on source strength; gamma ray irradiation imparts the masks with a faint odor (result from this study). |
| Electron beam | High, tunable beam current and dose rate<br>Widely used in pharmaceutical sterilization | High doses of electron radiation can degrade some plastics, difficult to achieve sufficiently high dose for sterilization using hospital equipment, low range of electrons forces an inline process limiting throughput |
| UV sterilization | UV sources are ubiquitous, inexpensive | Unclear if UV sterilizes throughout the thickness of the mask; requires line of sight; masks must be removed from biosafety container and handled individually |
| Dry heating<br>Steam heating | Ovens are cheap and plentiful | Many materials are temperature sensitive and not all materials in N95 masks are safe for heating; masks must be removed from biosafety container and handled individually |
| Vaporized Hydrogen Peroxide | Shown to work well, possibilities for batch processing | Tarnishes metal components; masks must be removed from biosafety container and handled individually |
| EtO (ethylene oxide) | Used widely in healthcare sterilization | Tarnishes metal components; limited throughput |
| Microwave oven | Quick, cheap, and easy | Melted and destroyed tested mask models |

**Table 2.** Number of masks of each type tested.

| Mask Type | Dose (kGy) |  |  |
| --- | --- | --- | --- |
|  | 0 | 10 | 50 |
| 3M 9105 | 3 | 3 | 3 |
| 3M 8210 | 3 | 3 | 3 + 1<sup>a</sup> |

<sup>a</sup> One mask in this category was improperly donned by the subject.

Both gamma and electron sources have been shown to accomplish viral inactivation in other arenas, such as food and mail sterilization. While the literature does not yet report on SARS-Cov2 itself, gamma ray radiation doses of between 5 and 10 kGy have been shown to achieve 4 log10 reduction of most viruses, including other single stranded RNA viruses [4, 11]. In particular, a 2019 study SARS-CoV was reported to be inactivated by a dose of 10 kGy, although lower doses were not studied [3]; and a 2015 study showed a 5 log10 reduction of MERS-CoV [10] with 10 kGy. Electron beam sterilization generally requires higher dose for the same inactivating effect [12, 13].

Gamma irradiation has a significant logistical advantage over other sterilization methods in that the soiled masks can be packaged and sealed in a container, brought to the gamma source, sterilized, and then removed without having to be unsealed (assuming the container could tolerate 10 kGy, and is not made of a high Z material). This could reduce the biosafety logistics surrounding sterilization as compared to UV, ovens, and chemical techniques. With the increasing evidence in recent days for the airborne transmission of SARS-CoV2, there is likely a need for biosafety level 3 management of soiled masks. Individual masks could also be bagged and labeled separately within the container.

^60^Co sources are often used in radiation therapy, food sterilization, and in basic science. Many hospitals across the world have access to ^60^Co sources as they are widely used for radiotherapy. However, ^60^Co sources decline in dose over time (half life = 5.6 years), so older units may not be able to provide a sufficient dose for mass sterilization. There are other sources for generating high energy gamma rays besides ^60^Co and our results on the operational survivability of the masks are applicable to any sterilization procedure with an equivalent dose and energy spectrum.

It should be emphasized that sterilization using gamma rays does not leave the masks radioactive. Specifically, the 1.17 and 1.33 MeV gamma rays emitted by ^60^Co as it decays will not induce secondary activation within the mask. This is in contradistinction to proton or neutron sources that could conceivably cause this effect.

## Methods

A set of five each of 3M 8210 and 9105 “duckbill” masks were irradiated to doses of 10 kGy and 50 kGy using a Gammacell 220 Excel Irradiator at MIT. The chamber in the irradiator is approximately 6 inches in diameter and 8 inches tall. Since the most radio-opaque component in the masks is a thin aluminum nose piece, which is still essentially transparent to the *∼*1.3 MeV photons produced in ^60^Co decay, the dose throughout the chamber should be fairly uniform.

The masks received either 10 kGy or 50 kGy of radiation emitted from the pencil rods in a circular pattern, at a dose rate of 2.2 kGy per hour. Dose variability was *≤*10%, based on previous characterization of this source. Spacers were used to position the masks in the most uniform flux region — if the chamber were fully packed the dose variability would be closer to 15%, and approximately 100 masks could be fit in the chamber.

**Fig. 1.**
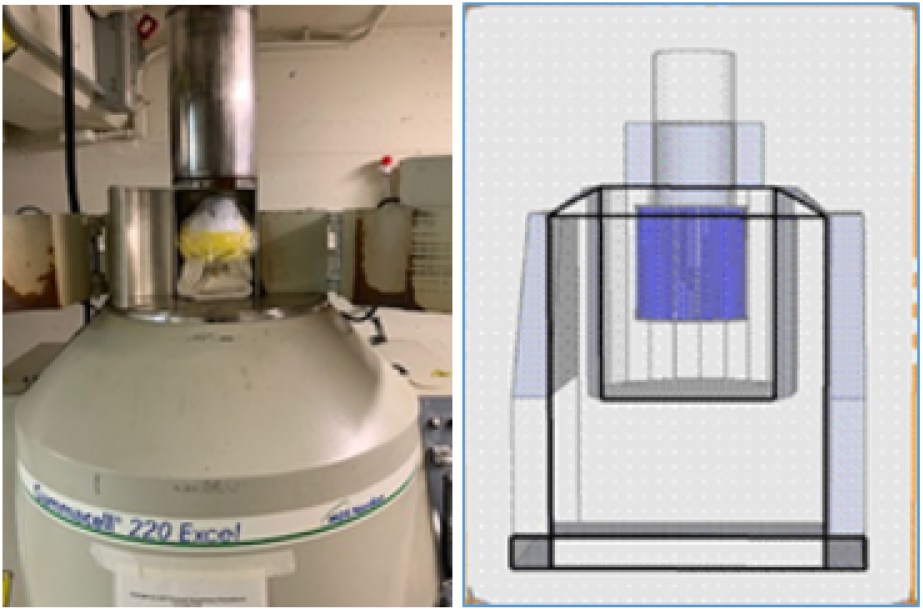
(left) Image of the Gammcell 220 Excel Irradiator with masks in the chamber (right) Cutaway diagram of the irradiator.

The OSHA-approved Gerson Qualitative Fit Test [7] QLFT 50 (saccharin apparatus) was utilized at Brigham and Women’s Hospital to check whether the masks suffered degradation during irradiation. Three masks of each type were tested at each dose level; all tests were performed blind on the subject. The test subject (M. P. S.) was first checked for taste sensitivity to saccharin using a dilute saccharin mist (eight sprays were sufficient to induce a clear taste sensation). Then, the fit test was repeated for each mask, handed to the test subject who was facing away from the masks to help exclude detection bias from the results. Masks from the three dose categories were tested in a random order. One test had to be repeated due to improper donning of the mask.

The masks that were not used in the qualitative fit test were tested for their particulate filtering efficiency at MIT (masks used in the qualitative fit test were not brought to MIT in an attempt to adhere to social distancing guidelines).

The masks were inserted into a specialized air duct, and ambient particulate matter was flowed through the duct (through the filter), with a pressure differential of approximately 175 pascals at 0.4 m/s face velocity. 0.3, 0.5, and 1 *µ*m particles were tested, and the filtering efficiency was measured using an optical particle counter (Aerotrak 9306, TSI Inc.).

## Results

All masks, including the unirradiated controls, the 10 kGy, and the 50 kGy masks passed a qualitative fit test (no saccharin was detected by the subject), with the exception of one mask that had received 50 kGy that was donned improperly. For the one 50 kGy-irradiated 8210 mask that did result in saccharin taste sensation by the test subject, the cause was found to be poor donning of the mask (the subject reported cool air flowing in around his nose while breathing in during that particular test). This was the second mask tested in the series of 18, and this test failure was believed to be due to inexperience on the part of the test subject in correctly applying a tight fit to the mask. After more practice, an additional 50 kGy-irradiated 8210 mask was randomly inserted into the test matrix, and it along with all other masks resulted in no detection of the saccharin mist. The one anomalous test served to confirm that the test subject could strongly taste the saccharin mist if it entered the mask, either through the filter or around the mask periphery. The subject did notice that some irradiated masks had a slight unrecognizable odor-the Food and Drug Administration notes that gamma ray sterilization of foods can impart an odor as well [14].

**Fig. 2.**
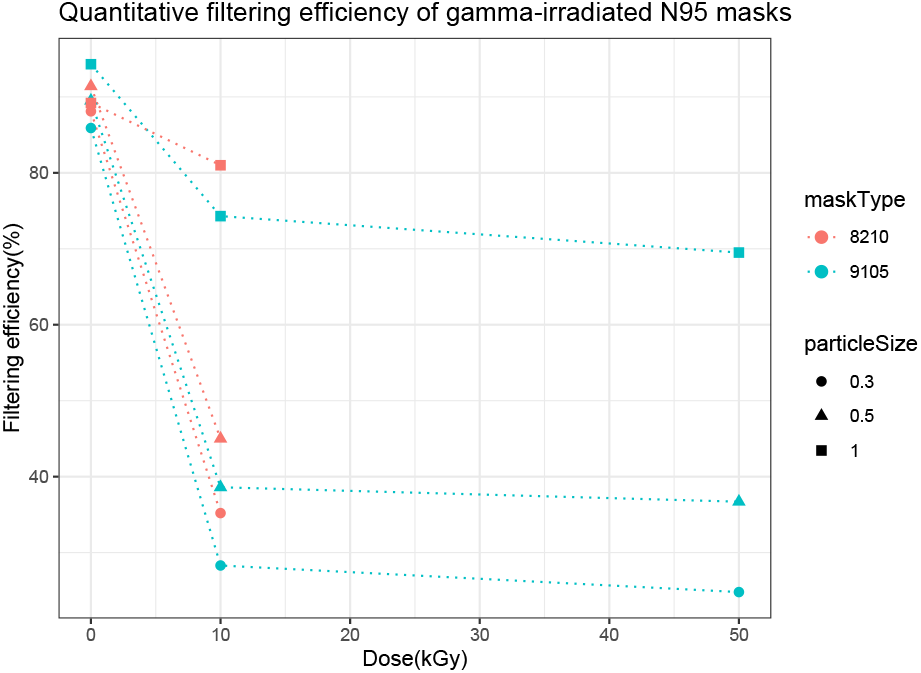
Filtering efficiency.

Particular matter filtration data are shown in table 3. The irradiated masks performed much more poorly than the unirradiated controls, especially in the filtration of 0.3 *µ*m particles.

**Table 3.**
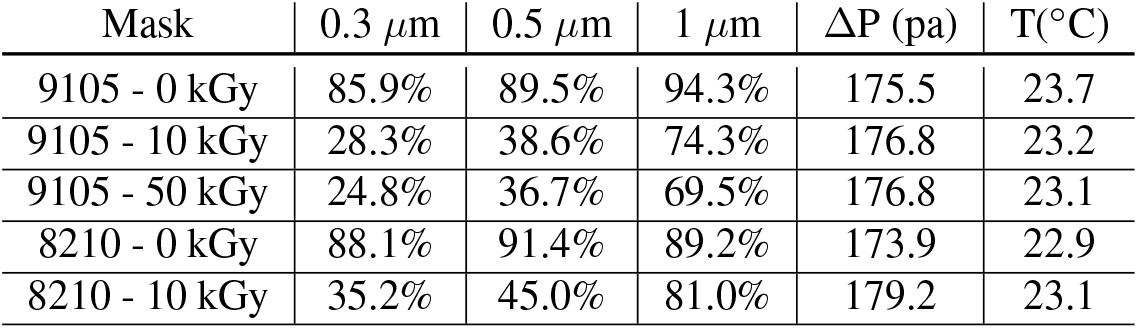
Ambient particulate matter filtration efficiency.

## Discussion

While disposable N95 masks remain functional as per an qualitative fit test after receiving both 10 kGy and 50 kGy doses of gamma radiation, the fact that their measured particular filtering efficiency declined so severely suggests against using ionizing radiation as a method of mask sterilization. The small particle filtration properties of a mask are provided by primarily electrostatic, rather than mechanical processes, so it is likely that the ionizing radiation is discharging the mask.

Furthermore, the fact that the masks continued to pass the qualitative fit test even after being irradiated is a significant finding, and suggests that the qualitative fit test alone cannot be used to assess the success or failure of an N95 sterilization procedure.

## Data Availability

All data, and specifics on the gamma source are available upon request.

## Acknowledgements

We would like to thank Profs. Michael H Lev, Anne White, Areg Danagoulian, Elazar Edelman, Alan Grossman, Zachary Hartwig, Dennis Whyte, Drs. Gordon Kose, Richard Lanza, William Bertozzi, Cody Dennett, Leigh Ann Kesler, John Boyer, as well as Julie Logan, Deborah Plana, Jose Gomez-Marquez, and Robert Hinshaw for their advice, support, and fruitful discussions.

## REFERENCES

[1] S. Feng, C. Shen, N. Xia, W. Song, M. Fan, and B. J. Cowling, “Rational use of face masks in the COVID-19 pandemic,” The Lancet Respiratory Medicine, p. S221326002030134X, Mar. 2020.

[2] D. J. Viscusi, M. S. Bergman, B. C. Eimer, and R. E. Shaffer, “Evaluation of Five Decontamination Methods for Filtering Facepiece Respirators,” Ann. Occ. Hyg., vol. 53, pp. 815–827, Oct. 2009.

[3] F. Feldmann, W. L. Shupert, E. Haddock, B. Twardoski, and H. Feldmann, “Gamma Irradiation as an Effective Method for Inactivation of Emerging Viral Pathogens,” The American Journal of Tropical Medicine and Hygiene, vol. 100, pp. 1275–1277, May 2019.

[4] R. Sullivan, A. C. Fassolitis, E. P. Larkin, R. B. Read, and J. T. Peeler, “Inactivation of Thirty Viruses by Gamma Radiation 1,” Applied Microbiology, vol. 22, no. 1, pp. 61–65, 1971.

[5] W. A. Rutala and D. J. Weber, “Disinfection and sterilization: An overview,” American Journal of Infection Control, vol. 41, pp. S2–S5, May 2013.

[6] C. R. Harrell, V. Djonov, C. Fellabaum, and V. Volarevic, “Risks of Using Sterilization by Gamma Radiation: The Other Side of the Coin,” International Journal of Medical Sciences, vol. 15, no. 3, pp. 274–279, 2018.

[7] “Fit Testing Procedures (Mandatory). - 1910.134 App A | Occupational Safety and Health Administration.”

[8] E. Joseph and E. Levenson, “NYC coronavirus: Hospitals could run out of needed supplies by next week, De Blasio says - CNN,” Mar. 2020.

[9] H. Bauchner, P. B. Fontanarosa, and E. H. Livingston, “Conserving Supply of Personal Protective Equipment—A Call for Ideas,” JAMA, Mar. 2020.

[10] M. Kumar, S. Mazur, B. L. Ork, E. Postnikova, L. E. Hensley, P. B. Jahrling, R. Johnson, and M. R. Holbrook, “Inactivation and safety testing of Middle East Respiratory Syndrome Coronavirus,” Journal of Virological Methods, vol. 223, pp. 13–18, Oct. 2015.

[11] F. C. Thomas, A. G. Davies, G. C. Dulac, N. G. Willis, G. Papp-Vid, and A. Girard, “Gamma Ray Inactivation of Some Animal Viruses,” Can. J. comp. Med, vol. 45, pp. 397–399, Oct. 1981.

[12] M. Plavsic and R. W. Nims, “Efficacy of Electron Beam for Viral Inactivation,” Journal of Microbial & Biochemical Technology, vol. 07, no. 04, 2015.

[13] G. Sanglay, Inactivation and Mechanism of Electron Beam Irradiation and Sodium Hypochlorite Sanitizers against a Human Norovirus Surrogate. PhD thesis, The Ohio State University, 2012.

[14] K. Morehouse and V. Komolprasert, “Overview of Irradiation of Food and Packaging,” in Irradiation of Food and Packaging, pp. 1–11, American Chemical Society, 2004.

